# Notable transmitted HIV drug resistance among people who inject drugs in Pakistan

**DOI:** 10.1101/2024.04.30.24306644

**Authors:** Stephanie Melnychuk, Laura H. Thompson, Chris Archibald, James F. Blanchard, Faran Emmanuel, Tahira Reza, Nosheen Dar, Paul Sandstrom, Souradet Y. Shaw, Marissa L. Becker, François Cholette

**Author notes:** Corresponding author: (FC).

## Abstract

Transmission of drug-resistant HIV strains to treatment-naïve patients can compromise antiretroviral therapy (ART) effectiveness and lead to treatment failure. In Pakistan, transmitted HIV drug resistance among people who inject drugs (PWID) is fuelled by a lack of ART, poor drug adherence, and unsafe injection practices, resulting in efficient transmission in large injecting networks. A cross-sectional study was conducted among PWID recruited in the Pakistani cities of Karachi, Larkana, Peshawar, Quetta and Hyderabad (August 2014 to January 2015). A portion of the HIV *pol* gene was amplified from HIV-reactive dried blood spot specimens (*n*=282/367) and sequenced using an in-house Sanger sequencing assay for HIV drug resistance mutation genotyping. Drug resistance mutations (DRMs) were identified using the Stanford University HIV Drug Resistance Database HIVdb algorithm (https://hivdb.stanford.edu/hivdb). Overall, HIV subtype A1 was dominant (78.0%; *n*=220), followed by CRF02_AG (15.6%; *n*=44), CRF35_AD (2.5% *n*=7), recombinants (3.5%; *n*=10), and subtype C (0.4% *n*=1). DRM analysis identified over half (63.8%) of participants harbored at least one DRM, of which 28.9% reported using help from a professional injector. Nearly all (99.4%) participants were not actively receiving ART because most (88.7%) had never undergone HIV testing and were unaware of their status. Findings suggest significant transmitted HIV drug resistance present among PWID, exacerbated by unsafe injection practices, particularly professional injection. Low testing rates signal a need for more comprehensive testing programs to improve HIV status awareness and ART coverage in Pakistan. Given most treatment-naïve participants had evidence of drug resistance, drug resistance genotyping prior to ART initiation might aid in ensuring effective treatment to prevent transmission of resistant HIV strains.

## Introduction

In Pakistan, HIV prevalence is estimated to be 0.1% in the general population, but is known to be much higher among people who inject drugs (PWID), with estimates as high as 40% of PWID in the country living with HIV (1,2). This considerable burden is attributable to the marginalization of PWID due to contextual factors, including illiteracy, poverty, homelessness, and criminalization (3–5), which constrain access to both harm reduction programs and antiretroviral therapy (ART) (6). Further, the requirement of individuals actively injecting drugs to undergo a two-week drug detoxification prior to ART initiation (7) and lack of a national opioid substitution therapy program in Pakistan presents structural barriers which might contribute to higher rates of HIV transmission among PWID overall, hindering effective HIV management and posing challenges to overall HIV epidemic control efforts in the country (8–10).

PWID undergoing ART have increased vulnerability to treatment interruptions due to a lack of social support and stable housing, mental health comorbidities, and effects of stigma and discrimination (9,11,12). Suboptimal therapy and non-adherence to ART facilitates the selection of viral strains harboring drug resistance mutations (DRMs) (13) which can be transmitted to ART-naïve individuals, constituting what is defined as transmitted drug resistance (14).

Transmitted drug resistance poses challenges to virological suppression and achieving undetectable viral loads in ART-naïve patients harboring DRMS (15,16), with studies documenting high risk of failing first-line therapy in ART-naïve patients harboring DRMs (17–19).

The disproportionate burden of transmitted drug resistance facing PWID may be fuelled by the rapid and efficient transmission of drug-resistant HIV strains in large, interconnected injecting networks with high frequency of sharing injecting equipment (20,21) in certain contexts, including Pakistan (22). High daily injection frequency (23), sharing syringes and other drug use paraphernalia (22), injection of shared blood-drug mixtures (24), and help from professional injectors (3), have been previously identified as risk factors among PWID in Pakistan.

Few studies have looked at baseline HIV DRMs among people living with HIV in Pakistan (5,25–27), with even fewer looking specifically at PWID. Knowledge of DRMs is essential for guiding ART selection, as ARTs are currently prescribed in Pakistan without baseline drug resistance genotyping (26). This increases the risk of ineffective treatment. Using data from a cross-sectional survey conducted in 2014 (28), we sought to assess HIV drug resistance in PWID across five major cities in Pakistan.

## Material and Methods

### Study setting

For this cross-sectional study, PWID were recruited from August 2014 to January 2015 across five major cities in Pakistan. To ensure accurate reflection the study population, a detailed mapping of size, geography, and operation typology of the PWID population was conducted prior to the survey as described previously elsewhere (29).

Following informed consent, participants were interviewed to collect information pertaining to sociodemographics, drug use practices, and utilization of HIV prevention and care services, as well as a dried blood spot (DBS) specimen.

### HIV serology and *pol* sequencing

DBS were shipped to the National Sexually Transmitted and Blood Borne Infection Laboratory (Public Health Agency of Canada, Winnipeg, Canada). Serological testing using the AVIOQ HIV Microelisa System (Avioq Inc., Durham, NC) was performed according to the manufacturer’s instructions. HIV drug resistance mutation genotyping was attempted on all HIV-reactive DBS.

A routine, in-house HIV drug resistance mutation genotyping assay was used to amplify and sequence a portion of the *pol* open reading frame (nt 2,074–3,334 on HXB2, <u>K03455</u>) from DBS as previously described (28). Sequence analysis and contig assembly was performed using RECall (30). HIV *pol* sequences are available via a public repository (i.e. NCBI) under accession numbers <u>MN887780</u>-<u>MN888069</u>.

### HIV subtyping and drug resistance mutation analysis

PWID HIV *pol* sequences were aligned, visually inspected, and manually edited in MEGA v7 (31) as required. Subtyping was carried out using REGA HIV automated subtyping tool v3 (32) and COMET (33), with SCUEAL (34) used as a tiebreaker for discordant results.

The Stanford HIV Drug Resistance Database program v9.4.1 (https://hivdb.stanford.edu) (35) was used to assess for mutations in the HIV *pol* gene associated with resistance to protease and reverse transcriptase inhibitors. Sequences containing one or more DRMs were considered drug-resistant (DR). Any DRMs identified for which no clinical implications were found in the literature were discarded.

### Statistical analyses

Descriptive statistics were calculated for sociodemographic and behavioral information. Pearson’s chi-square test was applied between DR classification and predictor variables using SciPy (36), where *P*<0.050 was used to define statistically significant associations.

### Ethics

Ethics board approval was obtained from the Health Research Ethics Board at the University of Manitoba [HS15691(H2012:294)] and BRIDGE Consultants Foundation, Pakistan. Informed consent was obtained verbally from study participants and documented by a member of the data collection team for the behavioural survey and biological sampling components of this study. Verbal consent was chosen due to varying levels of literacy among study participants and as an additional measure of protecting their identities.

## Results

### Study population and sequence dataset

A total of 1,453 PWID consented to participate in the study from Karachi (*n* = 300), Larkana (*n* = 300), Peshawar (*n* = 253), Quetta (*n* = 300) and Hyderabad (*n* = 300). All participants consented to providing a DBS sample. Serological testing identified 367 (25.5%) PWID living with HIV.

Of the HIV-reactive DBS specimens, 287 *pol* sequences (79.0%) were amplified – corresponding to 80% of HIV-reactive DBS from each city except Hyderabad, where amplification success was closer to 70%. In accordance with WHO HIV drug resistance criteria (37), five sequences containing premature stop codons were removed from subsequent analysis. Of 282 *pol* sequences, 109 (38.7%) were from Karachi, 90 (31.9%) from Peshawar, 39 (13.8%) from Hyderabad, 25 (8.9%) from Larkana, and 19 (6.7%) from Quetta. Most sequences were classified as subtype A1 (*n*=220, 78.0%), followed by CRF02_AG (*n*=44, 15.6%), CRF35_AD (*n*=7, 2.5%), recombinants (*n*=10, 3.5%), and subtype C (*n*=1, 0.4%).

### Sociodemographic characteristics

Sociodemographic and behavioral information of PWID for whom HIV sequences were obtained is presented in Table 1. All participants were male. Of 282 sequences analyzed, 180 (63.8%) were considered DR.

**Table 1.** Sociodemographic and behavioral characteristics of people who inject drugs in Pakistan stratified according to presence of drug resistance mutations in HIV sequences.

| Variable | DR ($\geq 1$ DRM)<br>($N=180$) | Non-DR<br>($N=102$) | $P^*$ |
| --- | --- | --- | --- |
| City, $n$ | | | $<0.001$ |
| Hyderabad | 35 | 4 |  |
| Karachi | 75 | 34 |  |
| Larkana | 22 | 3 |  |
| Peshawar | 30 | 60 |  |
| Quetta | 18 | 1 |  |
| Age, $n$ (%) | | | 0.010 |
| <24 | 43 (23.9) | 40 (39.2) |  |
| 25-29 | 71 (39.4) | 26 (25.5) |  |
| $\geq 30$ | 66 (36.7) | 36 (35.3) | |
| Years injecting drugs, $n$ (%)<sup>a</sup> | | | 0.200 |
| <2 | 32 (17.8) | 21 (20.6) |  |
| 2-4 | 64 (35.6) | 44 (43.1) |  |
| ≥5 | 84 (46.7) | 36 (35.3) |  |
| Times injecting the previous day, *n* (%) |  |  | 0.130 |
| 0 | 45 (25.0) | 15 (14.7) |  |
| 1 | 74 (41.1) | 48 (47.1) |  |
| ≥2 | 61 (31.7) | 39 (37.3) |  |
| Injection method at last injection, *n* (%) |  |  | 0.200 |
| Without help | 104 (57.8) | 70 (68.6) |  |
| Uses help, but not from a professional injector | 24 (13.3) | 10 (9.8) |  |
| Uses help from a professional injector | 52 (28.9) | 22 (21.6) |  |
| Current ART treatment status, *n* (%) |  |  | 0.720 |
| No | 19 (10.6) | 12 (11.8) |  |
| Yes | 1 (0.6) | 0 (0.0) |  |
| Not specified<sup>b</sup> | 160 (88.9) | 90 (88.2) |  |

\**P*-value based on Pearson chi-square test.
<sup>a</sup>Among respondents does not add to 100%.
<sup>b</sup>Low response rate for HIV treatment is because most participants had never undergone HIV testing and did not respond to questions regarding ART.

Statistically significant predictors of DR classification were city of residence and age (*P*<0.05) (Table 1). Years injecting, daily injection frequency, injection method, and ART status were not significantly associated with DR. Prevalence of DR ranged from 33.3% to 94.7% among participants when stratified by city. Of those with DR sequences, the highest proportion (39.4%) were between 25-29 years of age. Using help from a professional injector was slightly higher in DR participants (28.9%). Of participants who had undergone HIV testing and were aware of their positive status (*n*=32, 11.3%), one (3.1%) was actively receiving ART.

### Analysis of HIV drug resistance analysis

HIV mutations associated with drug resistance are summarized in Table 2. Of 282 analysed HIV *pol* sequences, 13 DRMs were identified among the 180 DR participants: two protease inhibitor (PI), two nucleoside RT inhibitor (NRTI), and nine non-nucleoside RT inhibitor (NNRTI) mutations (Table 2).

**Table 2.**
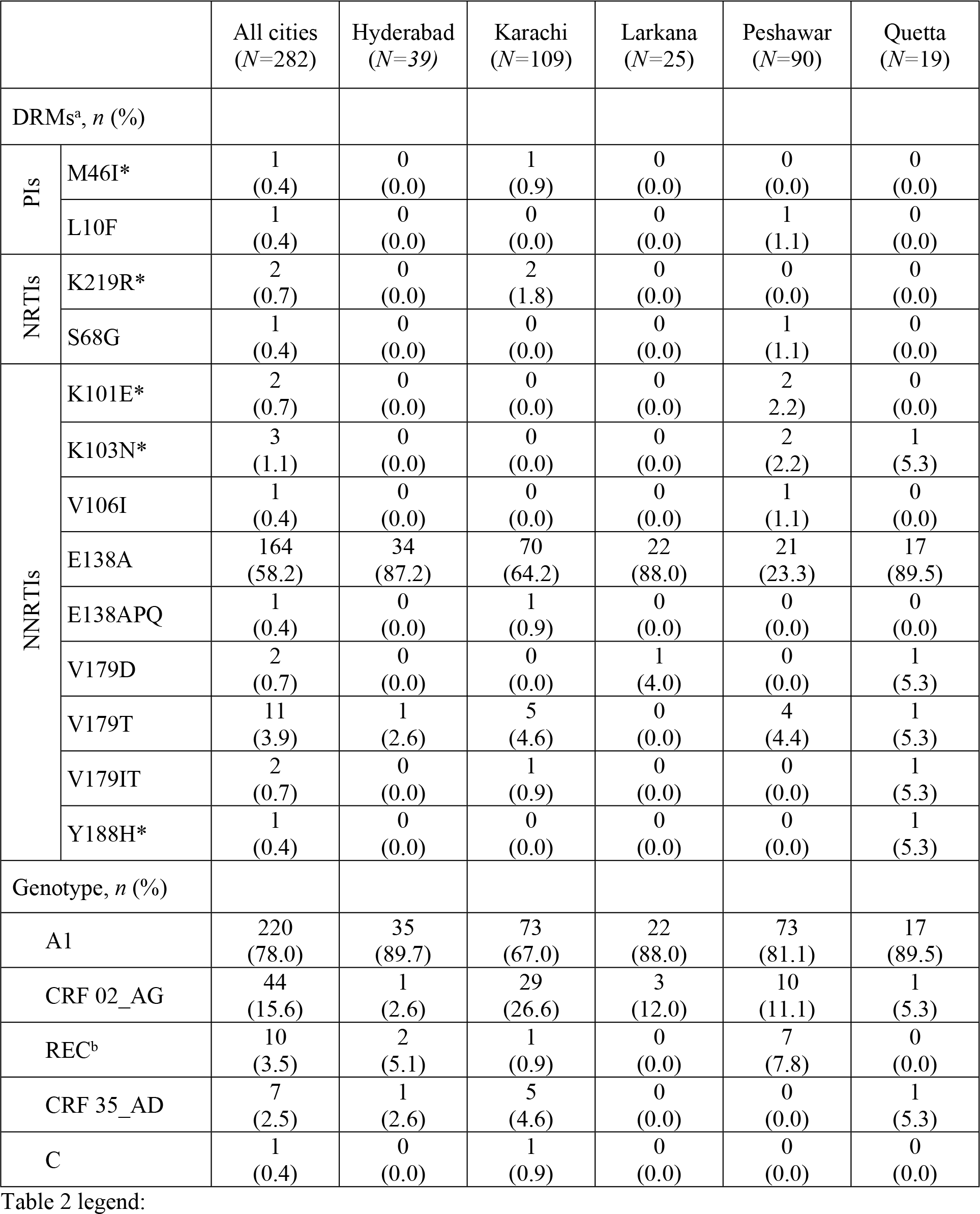
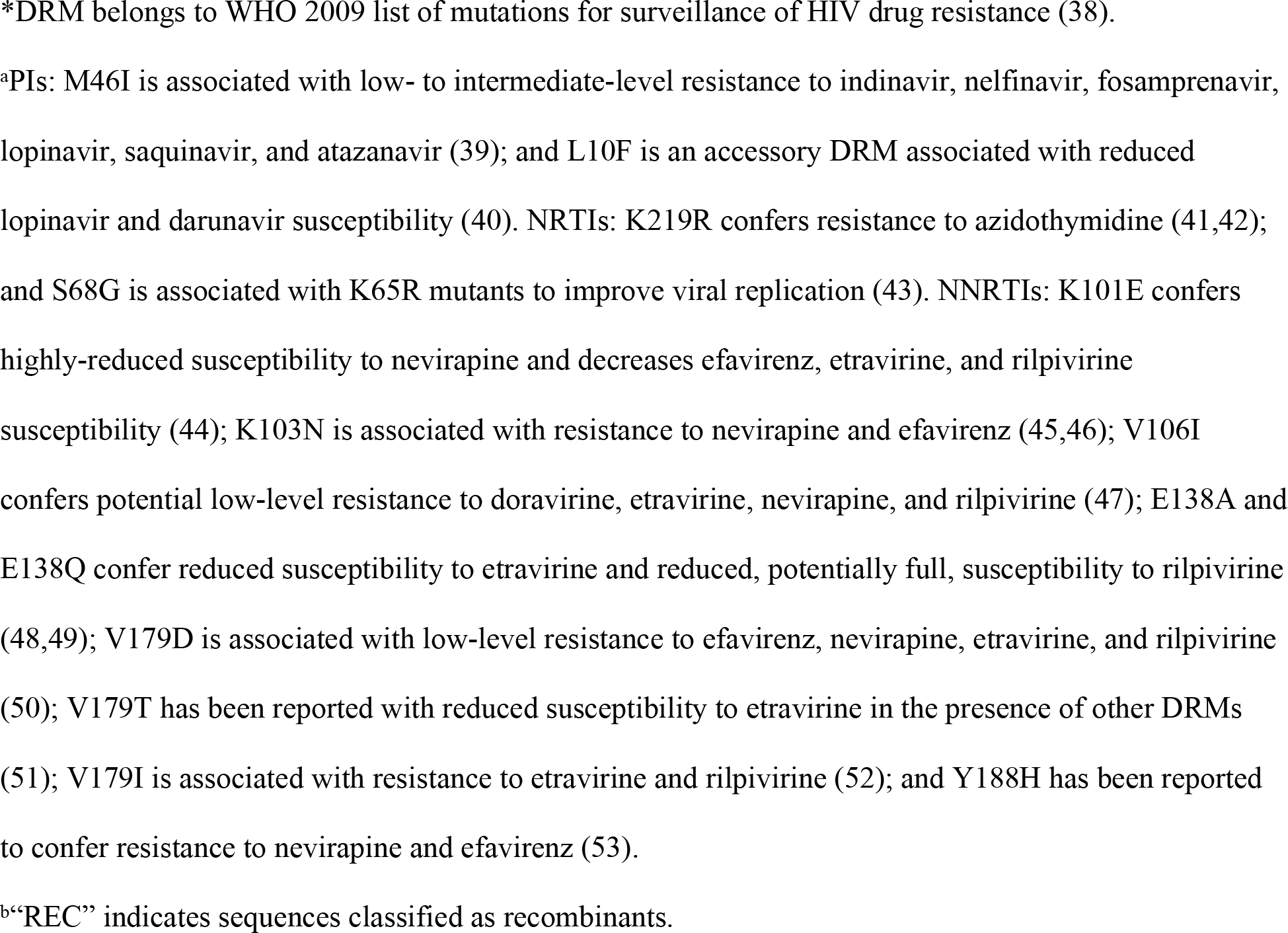
Drug-resistance mutations in HIV sequences from people who inject drugs across several Pakistani cities.

DRM E138A was the most common mutation across all cities (58.2%), with almost all sequences from Hyderabad, Larkana, and Quetta containing this mutation (87.2%, 88.0%, and 89.5%, respectively). Karachi sequences had the only reports of DRMs M46I, K219R, and V179T. Despite Peshawar having the smallest percentage (33.3%) of sequences containing any DRMs, these sequences had the most diversity, containing 7 of the 13 DRMs in our analysis, including the only observances of L10F, S68G, K101E, and V106I. The mutation K103N was only reported in two sequences from Peshawar and one sequence from Quetta.

## Discussion

Here, we present HIV drug resistance mutations from a cross-sectional survey conducted in 2014 among PWID from five cities in Pakistan. Our analysis demonstrated a high prevalence of DRMs associated with NNRTI resistance. The vast majority of participant sequences from Hyderabad (89.7%), Karachi (68.8%), Larkana (88.0%), and Quetta (94.7%) contained at least one DRM. Given that only one (0.6%) study participant reported actively receiving ART, observing DRMs in high frequency among participants suggests significant HIV transmitted drug resistance present among PWID in Pakistan.

Over half (58.2%) of PWID in our study harbored E138A, conferring low-level resistance to the NNRTIs etravirine and rilpivirine (48,49). Other studies have observed E138A among people living with HIV in Pakistan, reporting varying prevalence in ART-experienced (13-78%) and ART-naïve (8-39%) cohorts (5,27). Perhaps more concerning are the DRMs observed in low prevalence conferring resistance to efavirenz– namely K101E (44), K103N (45,46), V179D (50), and V188H (53). Pakistan’s first-line ART regimen for adults is currently tenofovir and lamivudine with dolutegravir (54), although efavirenz was formerly used in first-line regimes at the time of our study (55) and is currently suggested for use in alternate regimes. Our results suggest first-line regimes would be effective treatment for most PWID living with HIV who participated in our study, although this might not be true for previous regimes. While Pakistan has second-line regimens, these drugs are more prone to shortages compared with first-line regimes (56), posing challenges to HIV treatment in individuals harboring resistance to efavirenz prior to the change to dolutegravir in 2019 (57) and enabling transmission of drug-resistant strains.

We observed city of residence and age to be significantly associated with DR classification. Participants with DR sequences reported slightly higher prevalence of help from a professional injector compared to non-DR sequences (28.9% versus 21.6% respectively), and while method at last injection was not significantly associated to DR, we did not stratify the analysis by city and injection method to determine its significance for each study city.

Professional injectors (or street doctor) are PWID themselves who reuse the same injecting equipment to inject several clients – often younger and inexperienced PWID – daily, for a fee (58–60). This fee can be something known as “scale”, a blood-drug mixture collected by the professional injector by double-pumping the syringe, which is then sold to other PWID for less than the regular drug (22,28,61), allowing for efficient transmission of drug-resistant HIV. Using help from professional injectors has previously been reported to be more prevalent in Karachi (53.7%) compared to Peshawar (25.2%) and Quetta (19.6%) (59). Differences in DRM prevalence among cities might also be partially explained by varying rates of heroin injection, observed to be high in Karachi and Quetta (96.9% and 81.1% respectively) (59). Heroin has been associated with high-risk injection practices, such as needle sharing and “jerking”, where heroin is mixed with blood and shared amongst injectors (6), facilitating transmission of resistant strains among PWID.

In this study, we report the spread of drug-resistant HIV is likely being fuelled by the lack of ART coverage among PWID participants, permitting high viral loads which enable efficient HIV transmission. Our findings stress the lack of ART coverage among PWID in Pakistan, known to still be an ongoing issue and major obstacle to HIV treatment in the country. The low ART coverage observed in our study is likely due in part to the lack of participants’ awareness that they are living with HIV overall, as we observed nearly all (88.7%) PWID participants to have never undergone HIV testing. Awareness of HIV status among people in Pakistan is known to be low (21%), with people living with HIV actively receiving ART being even lower (12%) (62). Moreover, it is well established that PWID face barriers to ART access, specifically identifying Pakistan’s lack of national opioid substitution therapy program as a key intervention to HIV prevention that remains illegal (8,11).

## Conclusions

By sequencing, genotyping, and assessing for HIV drug resistance, we have been able to identify the circulating HIV strains and the level of drug resistance among PWID in Pakistan.

Overall, our study indicates that few DRMs are found amongst PWID in Pakistan conferring low-level resistance to etravirine, rilpivirine, and efavirenz. While our results suggest the majority of PWID in our study would be susceptible to current first-line ART regimes, perhaps more concerning is their lack of awareness of HIV-positive status and extremely limited access to treatment. These findings highlight a need for more comprehensive testing programs among PWID to increase HIV status awareness and drug resistance genotyping prior to ART initiation to ensure effective treatment of resistant HIV strains.

## Data Availability

The data that support the findings of this study have been deposited in a public repository (i.e. NCBI) under accession numbers MN887780-MN888069.

## Acknowledgements

We thank the surveillance team for their efforts in mapping, data collection, specimen collection and data entry. We also thank the participants who took part in the surveys for their time and participation. We thank Kiana Kadivar (National Sexually Transmitted and Blood Borne Infection Laboratory) for her initial assistance with coordinating serological testing.

## Notes

### Competing Interest Statement

The authors have declared no competing interest.

### Funding Statement

The author(s) received no specific funding for this work.

